# Spatial covariance analysis of FDG-PET and HMPAO-SPECT for the differential diagnosis of dementia with Lewy bodies and Alzheimer’s disease

**DOI:** 10.1101/2021.10.26.21265551

**Authors:** Matthew Ingram, Sean J. Colloby, Michael J. Firbank, Jim J. Lloyd, John T. O’Brien, John-Paul Taylor

**Author notes:** **Corresponding author** Matthew Ingram, Medical Education Unit, Princess Alexandra Hospital, 199 Ipswich Road, Woolloongabba, QLD 4102, Australia.

## Abstract

We investigated diagnostic characteristics of spatial covariance analysis (SCA) of FDG-PET and HMPAO-SPECT scans in the differential diagnosis of dementia with Lewy bodies (DLB) and Alzheimer’s disease (AD), in comparison with visual ratings and region of interest (ROI) analysis. Sixty-seven patients (DLB 29, AD 38) had both HMPAO-SPECT and FDG-PET scans. Spatial covariance patterns were used to separate AD and DLB in an initial derivation group (DLB n=15, AD n=19), before being forward applied to an independent group (DLB n=14, AD n=19). Visual ratings were by consensus, with ROI analysis utilising medial occipital/medial temporal uptake ratios. SCA of HMPAO-SPECT performed poorly (AUC 0.59±0.10), whilst SCA of FDG-PET (AUC 0.83±0.07) was significantly better. For FDG-PET, SCA showed similar diagnostic performance to ROI analysis (AUC 0.84±0.08) and visual rating (AUC 0.82±0.08). In contrast to ROI analysis, there was little concordance between SCA and visual ratings of FDG-PET scans. We conclude that SCA of FDG-PET outperforms that of HMPAO-SPECT and performed similarly to other analytical approaches, with the potential to improve with larger derivation groups. Compared to visual rating, SCA of FDG-PET relies on different sources of group variance to separate DLB from AD.

## 1. Introduction

Dementia with Lewy bodies (DLB) is a common form of late life degenerative dementia.^1, 2^ The differential diagnosis of DLB can be challenging, as many clinical features, such as cognitive fluctuations, subtle motor signs and rapid-eye movement (REM) sleep disorder, can be difficult to recognise clinically.^3, 4^ The difficulty is compounded by the possibility of Alzheimer’s disease (AD) co-pathology influencing the clinical phenotype.^4-6^ Consequentially, whilst post-mortem studies indicate that DLB accounts for over 15% of all dementia cases, the prevalence of DLB clinically identified antemortem is just 4-5%.^1, 7^ Identifying patients with DLB therefore remains important, both for future research and for patient prognosis and management, which is clearly different in DLB compared to other dementias like AD.^7-9^

Numerous biomarkers have been proposed to aid clinicians in identifying DLB, of which ^123^I-FP-CIT SPECT (DaTScan) and ^123^I-MIBG cardiac scintigraphy are the best supported, as reflected by their recommendation as indicative biomarkers in the recent DLB consensus criteria and some national guidelines.^2, 10-13^ FDG-PET and HMPAO-SPECT were suggested as supportive biomarkers according to the same criteria.^2^ Whilst not as well evidenced as the indicative biomarkers for DLB, both FDG-PET and HMPAO-SPECT are indirect surrogates of neuronal activity, and by extension neuronal integrity, features that are influenced by a variety of different pathologies.^14, 15^ They can offer broad clinical diagnostic utility of the different subtypes of dementia, such as frontotemporal dementia and Alzheimer’s disease dementia (in contrast to ^123^I-FP-CIT SPECT, which is largely specific to a diagnosis of DLB vs. other dementias).^11^ Additionally, both are more widely available than ^123^I-FP-CIT SPECT.^15,16^ Enhancing their utility in distinguishing DLB would therefore be of added benefit clinically, both in terms of reducing the need for additional expensive and radiation associated investigations, as well as picking up on DLB cases which may not be apparent clinically.

Current practice is for visual interpretation of these scans, but this is prone to a high degree of inter-rater variation.^17, 18^ Quantification methods, such as region of interest (ROI), have thus been proposed and, in both FDG-PET and HMPAO-SPECT, have achieved similar or better accuracy than visual rating for the diagnosis of DLB.^19, 20^ Most of these techniques are univariate, quantifying tracer uptake within individual brain regions or voxels and treating them as independent variables.^20, 21^ These are able to identify established DLB-specific features, such as preserved posterior cingulate uptake on a background of occipital hypometabolism.^21^ However, these features are also used to guide visual interpretation.^2, 22^ Therefore, whilst univariate methods may offer improved reliability in detecting observable changes, they seem unlikely to identify DLB where recognised visual cues are absent.

Alternative multivariate approaches such as spatial covariance analysis (SCA),^23^ an extension of principal component analysis (PCA), examine the inter-relationships of tracer-uptake between voxels, potentially allowing the capture of additional sources of disease-group variance.^19, 23^ After identifying the covariance patterns of tracer uptake that best separate two disease groups, independent subjects can then be classified according to the degree in which they express these patterns.^23, 24^ SCA is particularly tractable in functional modalities, such as FDG-PET and HMPAO-SPECT, where disease-state has the potential to affect tracer uptake to widespread brain regions, and relationships in tracer uptake may represent physiologically meaningful characteristics, such as inter-regional activity.^19, 25^ SCA of HMPAO-SPECT data in the differential diagnosis of AD and DLB has previously demonstrated greater diagnostic utility when compared to visual rating and univariate ROI methods.^19^ Whilst HMAPO-SPECT remains cheaper and more accessible, it is well established that FDG-PET is superior ^2, 15, 20, 26^ and as a modality is also becoming more widely available, largely due to its role in oncology.^15^ Therefore, our primary aim was to compare the diagnostic performance of SCA between FDG-PET and HMPAO-SPECT, in the same group of patients. The secondary aim was to compare the diagnostic accuracy of SCA with those of visual rating and ROI analysis, in both imaging modalities. We also contrasted the relationships between SCA expression and ROI scores with visual ratings, in order to identify whether they rely on different sources of variation to distinguish between AD and DLB. This is of interest as identification of additional sources of image variance, distinct from those used during visual rating, could possibly improve differential diagnosis.

## 1. Methods

### 2.1 Patient recruitment

Analysis was carried out on AD and DLB groups reported in a previous paper, where HMPAO-SPECT and FDG-PET were compared in terms of visual ratings and ROI procedures.^20^ Patients were recruited from individuals aged over 60 years with mild to moderate dementia who had been referred to clinical services in North-East England. All participants met clinical criteria for either probable AD^27^ or probable DLB.^3^ Diagnosis was by consensus between 2 experienced clinicians; neither FDG-PET nor perfusion SPECT scans had been used to confirm the diagnosis^20^. Exclusion criteria were past history of alcohol or drug dependence, contraindications to FDG-PET or HMPAO-SPECT scanning, or fasting blood glucose levels greater than 180 mg/dL. In total, 38 AD and 29 DLB patients were randomly assigned to either a derivation (19 AD, 15 DLB) or independent (19 AD, 14 DLB) group. All subjects underwent FDG-PET and HMPAO-SPECT scanning.

Patients underwent clinical and cognitive assessments, including the Cambridge Cognitive Examination (CAMCOG),^28^ mini mental state examination (MMSE)^29^ and the motor subsection of the Unified Parkinson Disease Rating Scale (UPDRS III).^30^

Study approval was by Newcastle and North Tyneside 1 Research Ethics Committee (REF 09/H0906/88), and all participants (or nominated Independent Mental Capacity Advocate where participant lacked capacity) gave written informed consent.

### 2.2 Scanning

SPECT scans were acquired using a Siemens Symbia S dual-detector gamma camera, 30 minutes after the intravenous administration of 500 MBq of ^99m^Tc-HMPAO. One hundred and twenty 25 second planar views were obtained on a 128 × 128 matrix, zoom 1.23 (pixel size, 3.9 mm), using a low-energy high-resolution collimator and circular orbit with a typical radius of 14 cm. Section images were produced with Hybrid Recon Neurology software (Hermes Medical Solution Ltd.) via ordered-subset expectation maximization iterative reconstruction with 4 iterations and 20 subsets. They were post-filtered using a 3D Gaussian filter of 1.1 cm full width at half maximum (FWHM) and uniform attenuation correction. FDG-PET scans were obtained over 10 minutes using a Siemens Biograph Truepoint PET/CT scanner 30 minutes after intravenous administration of 250 MBq of ^18^F-FDG. Siemens software was used for iterative reconstruction, with scatter and attenuation correction based on CT scan data acquired immediately prior to the ^18^F-FDG PET scan. For all scans patients were injected in quiet surroundings with eyes open.

### 2.3 Spatial pre-processing

All images were spatially normalized using an affine transform (12 parameters) to match respective FDG-PET and HMPAO-SPECT templates in standard MNI (Montreal Neurological Institute; http://www.bic.mni.mcgill.ca/) space using FMRIB’s linear image registration tool (http://www.fmrib.ox.ac.uk/fsl/flirt/index.html). Generation of specific FDG-PET and HMPAO-SPECT templates have been previously described.^19, 20^ Images were then visually inspected to ensure accuracy of registrations. A 10mm FWHM 3D Gaussian filter was applied to the registered scans.

### 2.4 Multivariate spatial covariance analysis

Voxel-based PCA was separately applied to spatially pre-processed HMPAO-SPECT and FDG-PET images from the derivation sample (AD, n = 19; DLB, n = 15) using generalized covariance analysis software (http://www.nitrc.org/projects/gcva_pca/).^31^ This captured the major sources of between and within group variation. A SPECT/PET binary mask image defined the brain volume subspace for voxelwise analyses. This generated a series of PCA eigenimages in a decreasing order of variance. The voxels within each eigenimage had either positive or negative weights, where the weights express the strength of interaction between voxels contributing to the eigenimage. Voxels with positive and negative weighting respectively demonstrate concomitant increased and decreased tracer uptake. Once calculated, eigenimage weights were fixed and equal for all subjects. The degree to which subject_i_ expressed each eigenimage_j_ was represented by means of the subject scaling factor SSF_i, j_, which was calculated as the product between the corresponding voxel values in the pre-processed scan and eigenimage, summed over the brain volume subspace defined by the SPECT/PET binary mask image. A higher SSF score represented more concurrent increased tracer uptake to voxels with positive weights, and more concurrent decreased tracer uptake to voxels with negative weights. To identify the spatial covariance pattern (SCP) that best-discriminated DLB from AD, each individual’s SSF_i, j_ were entered into a linear regression model as predictor variables with group (AD, DLB) as the dependent measure. Akaike’s information criteria (AIC) determined the optimum number of eigenimages_j_ to be included in the model.^19^ For HMPAO-SPECT, eigenimages 4, 5 and 7 yielded the lowest AIC value and were selected as predictors for the regression model. Linear combination of these eigenimages generated the composite spatial covariance pattern (SCP_SPECT_) that optimally distinguished DLB from AD, accounting for up to 5.6% of the total image data variance. The degree to which each subject expressed the SCP_SPECT_ was characterised by the SSF_SPECT_. For FDG-PET, eigenimages 1, 4, 5, 9, 11 and 12 yielded the lowest AIC value and generated the SCP_PET_ that optimally differentiated DLB from AD, accounting for up to 29.4% of the total image data variance. The degree to which each subject expressed the SCP_PET_ was characterised by the SSF_PET_.

Stability of the voxel weights from the SCP_(SPECT)_ and SCP_(PET)_ scans were assessed using a bootstrap resampling procedure with 1000 iterations. This identified regions that significantly contributed to the patterns with high confidence. The voxel weights of each SCP are converted into Z maps, calculated at each voxel location as the ratio of voxel weight to corresponding bootstrap derived standard deviation. The Z-statistic follows approximately a standard normal distribution and a one-tailed p≤0.05 implies a threshold of |Z| ≥ 1.64.^32^

To investigate the diagnostic value of the SCP_SPECT_ and SCP_PET_ in differentiating between AD and DLB, the patterns were then tested on the pre-processed images from the independent group (AD, n = 19; DLB, n = 14). This was performed by operation of forward application, with every voxel in a subject scan being multiplied by the corresponding voxel weight in the SCP, then summed over the brain volume subspace defined by the SPECT/PET binary mask image. The resultant number denotes to what extent a subject expresses the model SCP. Using this technique, each individual’s SSF of the respective model SCP_SPECT_ and SCP_PET_ were computed (SSF_(SCP_SPECT)_, SSF_(SCP_PET)_).

For ease of presentation and comparison with results from ROI analysis, we linearly rescaled all derivation FDG-PET and HMPAO-SPECT SSF’s to yield the SCA score (range 0 to 1).

### 2.5 Visual analysis

Visual ratings of the FDG-PET and HMPAO-SPECT scans have been previously reported.^20^ In short, visual analysis was performed by 3 medical physicists. Two of the physicists were clinical scientists who were fully approved by their National Health Service (NHS) employer for diagnostic reporting of scans, with a minimum of 20 years of experience in diagnostic brain scan reporting in dementia within the NHS. The third physicist had 10 years of research experience in neuroimaging analysis in dementia. Reviewers were blind to clinical information; randomised identification labels were generated for both FDG-PET and HMPAO-SPECT scans, making comparison between matching 18F-FDG PET and HMPAO-SPECT scans impossible.^20^

Each reader independently rated each scan for a match to AD or DLB using a 5-point scale (Definitely AD, Probably AD, Unclear, Probably DLB, Definitely DLB). The reporting team then met to produce a set of consensus ratings for each scan, using the same scale, and to make a final diagnostic decision (DLB, AD or unclear).^20^ When comparing concordance of diagnostic decisions between quantitative analyses and visual analysis, we transformed the final visual diagnostic decision into a binary classifier of DLB (DLB on final decision) or not DLB (AD or unclear on final decision).

### 2.6 ROI method

The ROI results in these patients have also been previously reported.^20^ To create study and modality-specific templates, each SPECT scan was spatially coregistered with that subject’s 18F-FDG PET scan. The 18F-FDG PET scans were then spatially aligned the standard template in SPM8 (www.fil.ion.ucl.ac.uk/spm), with the same alignment parameters then being applied to the SPECT scans. Averaging the aligned scans gave the study-specific templates for SPECT and 18F-FDG PET data, to which the original scans were (nonlinearly) spatially normalized. Ratios in tracer uptake between a number of regions of interest (ROIs, from the AAL atlas^33^) were evaluated.^20^ As supported by previous work,^20, 34^ the ratio of tracer uptake in the medial occipital lobe relative to medial temporal lobe (MTL) gave the greatest separation between AD and DLB, and was calculated for each scan. For ease of comparison with SCA, the ROI ratios were then multiplied by -1 and linearly rescaled. As with SCA, the ROI scores ranged between 0 and 1.

### 2.7 Statistical analyses

Data were exported and statistical analyses carried out in the Statistical Package for Social Sciences (SPSS version 19.0, https://www.ibm.com/analytics/spss-statistics-software) and R (R version 4.0.2 for Windows, https://cran.r-project.org/bin/windows/base/index.html). Continuous variables were tested for normality of distribution using the Shapiro-Wilk test. Differences in demographic, clinical, and imaging variables were examined where appropriate using parametric (ANOVA) and nonparametric (Mann–Whitney U) tests.

Diagnostic characteristics of SCA scores, ROI and visual analysis (using consensus visual ratings) in distinguishing DLB from AD were determined using area under the curve (AUC) from receiver operating characteristic (ROC) analyses.^35^ Statistical significance of AUC differences were calculated ^36^ by the De-Long et al method.

Cohen’s kappa (κ) and percentage agreement were used to quantify concordance,^37, 38^ between the diagnostic decisions (DLB vs not DLB) from the two quantification methods and from visual analysis, for scans obtained from participants in the independent group. All statistical tests were reported as significant if P < 0.05.

## 2. Results

### 3.1 Participant demographics and clinical characteristics

Participant demographics and clinical scores are presented in Table 1. No statistically significant differences in demographics or clinical scores were apparent between the derivation and independent groups. In both groups, UPDRS scores were, as expected, significantly higher in DLB, whereas age, gender, MMSE and CAMCOG were similar between AD and DLB.

**Table 1.** Demographics and clinical characteristics of the derivation and independent groups. Values expressed as mean ± standard deviation. AD, Alzheimer’s disease; DLB, dementia with Lewy bodies; MMSE, mini mental state examination; CAMCOG, Cambridge Cognitive Examination; UPDRS, Unified Parkinson’s Disease Rating Scale. Significant (p < 0.05) results in bold.

|  | AD | DLB | P value |
| --- | --- | --- | --- |
| Derivation |  |  |  |
| n | 19 | 15 |  |
| Gender (m:f) | 11:8 | 11:4 | $\chi^2 = 0.875$, $p = 0.350$ |
| Age (years) | $75.6 \pm 8.2$ | $75.4 \pm 6.3$ | $F_{1,32} = 0.004$, $p = 0.949$ |
| MMSE (<sub>max</sub>30) | $21.1 \pm 3.5$ | $22.0 \pm 4.7$ | $U = 122.500$, $p = 0.493$ |
| CAMCOG (<sub>max</sub>107) | $72.7 \pm 11.8$ | $71.9 \pm 10.3$ | $U = 137.500$, $p = 0.864$ |
| UPDRS (<sub>max</sub>199) | $3.5 \pm 2.8$ | $24.7 \pm 12.2$ | **$U = 1.000$, $p < 0.001$** |
| Independent |  |  |  |
| n | 19 | 14 |  |
| Gender (m:f) | 11:8 | 11:3 | $\chi^2 = 1.551$, $p = 0.213$ |
| Age (years) | $76.0 \pm 6.9$ | $76.6 \pm 4.5$ | $F_{1,31} = 0.073$, $p = 0.788$ |
| MMSE (<sub>max</sub>30) | $20.8 \pm 3.9$ | $21.6 \pm 4.1$ | $U = 115.500$, $p = 0.529$ |
| CAMCOG (<sub>max</sub>107) | $70.0 \pm 11.5$ | $72.3 \pm 16.3$ | $U = 112.000$, $p = 0.461$ |
| UPDRS (<sub>max</sub>199) | $3.7 \pm 3.7$ | $24.8 \pm 12.2$ | **$U = 3.000$, $p < 0.001$** |

### 3.2 Spatial covariance analysis

The voxel SCPs determined with SCA in the derivation group, from HMPAO-SPECT and FDG-PET data, are shown in Figure 1. The patterns show areas of relative increased (red) and decreased (blue) tracer uptake, in the DLB group relative to the AD group. For HMPAO-SPECT there was concomitantly increased tracer uptake in the anterior and posterior cerebellum, cerebellar tonsil, medial frontal gyrus, inferior and superior temporal gyri, the uncus, amygdala, hippocampus and precuneus. There was concomitantly decreased uptake in the superior frontal, inferior occipital, precentral, lingual and middle occipital gyri. For FDG-PET, there was concomitantly increased uptake in hippocampus, amygdala, posterior cingulate, anterior cerebellum and the superior, inferior and middle temporal gyri with concomitantly decreased uptake in precuneus, inferior parietal lobe, lingual gyrus and middle and superior frontal gyri. Both SCPs show increased concomitant tracer uptake to the right temporal lobe; aside from this, however, there were relatively few similarities between the two discriminatory patterns. All regions that contributed significantly to the HMPAO-SPECT and FDG-PET patterns are shown in the Supplementary tables 1 and 2.

**Figure 1.**
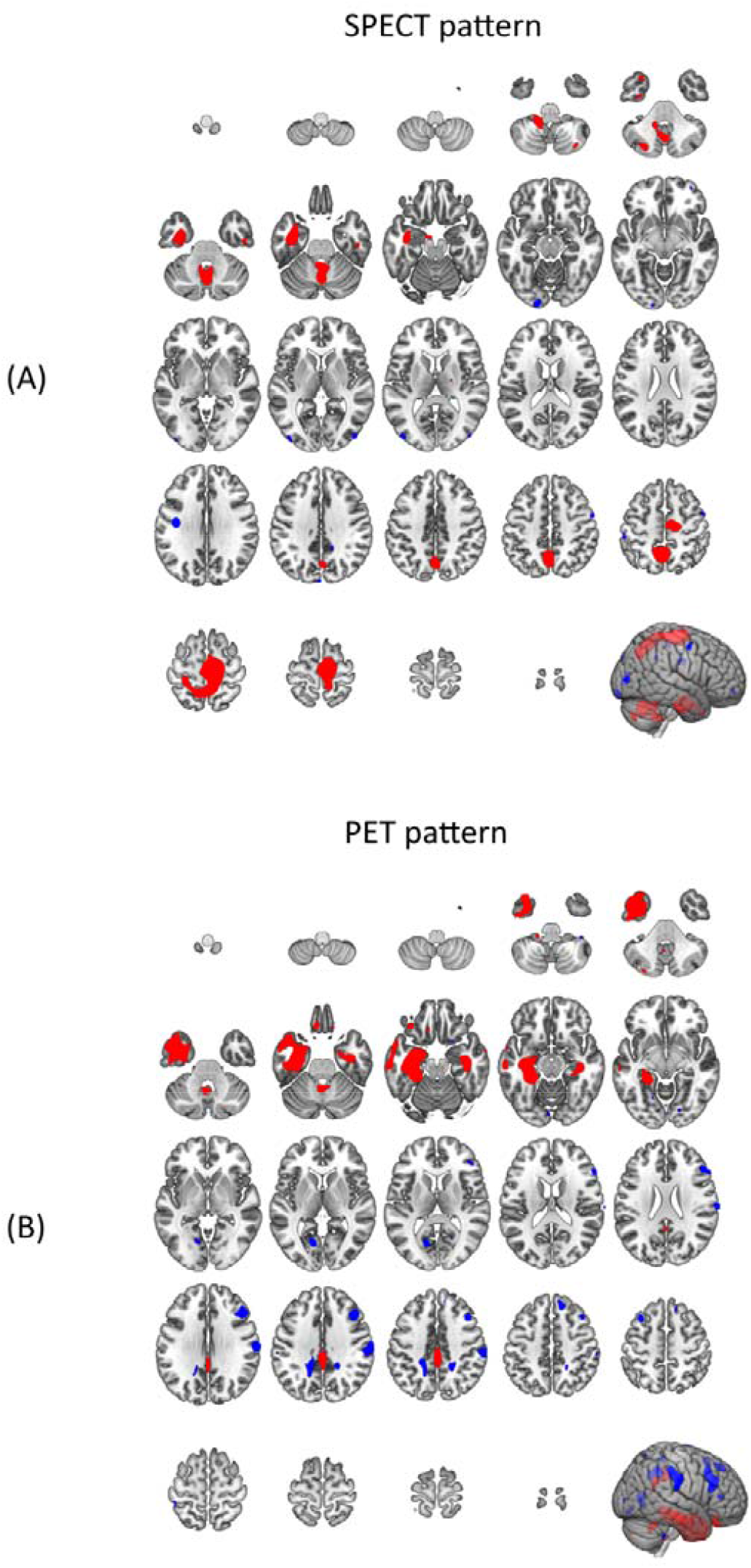
Spatial covariance patterns identified by HMPAO-SPECT (A) and FDG-PET (B), which were more strongly expressed in DLB than in AD. Red areas show regions of significant and concomitantly increased rCBF/FDG-uptake, blue areas show regions of significant concomitantly decreased rCBF/FDG-uptake. rCBF=regional cerebral blood flow.

### 3.3 SCA HMPAO-SPECT as a diagnostic tool

The SCA scores generated from HMPAO-SPECT scans from the derivation and independent groups are shown in Figure 2. Within the derivation group, patients with AD had significantly lower SCA scores than those with DLB (F_1,32_ = 17.6, p < 0.001; 0.288 ± 0.160 [AD mean ± s.d.], 0.569 ± 0.229 [DLB mean ± s.d.]). Using ROC curve analysis, the optimum threshold (dotted line) for separating the two groups showed very good diagnostic characteristics (AUC ± standard error, DLB sensitivity, AD specificity; 0.86±0.07, 80%, 79%). The optimum threshold was then applied to the independent group, where no significant differences were found in SCA scores between AD and DLB (U = 108.000, p = 0.189; AD 0.355 ± 0.151, DLB 0.435 ± 0.152), with poor diagnostic characteristics (AUC 0.59 ± 0.10, DLB sensitivity 50%, AD specificity 63%).

**Figure 2.**
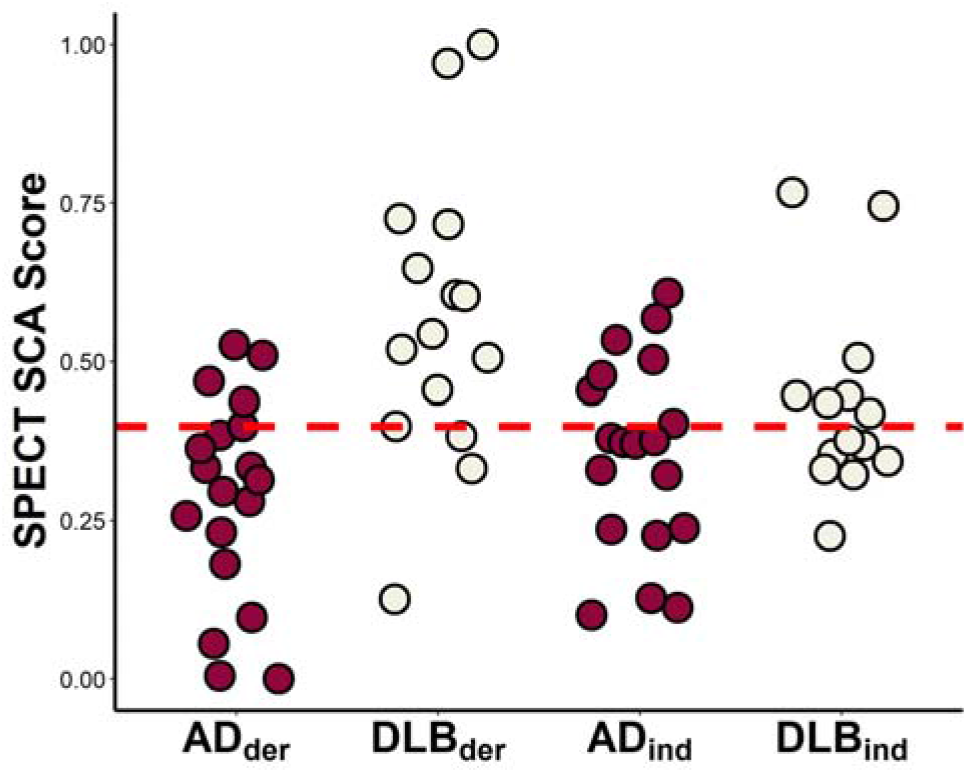
Distribution of SCA scores generated from HMPAO-SPECT scans, in the derivation (AD_der_ n=19, DLB_der_ n=15) and independent (AD_ind_ n=19, DLB_ind_ n=14) cohorts. Dotted line indicates separation threshold as calculated from the derivation cohort. Cases above the line were classified as DLB, below as AD. SCA, spatial covariance analysis; AD, Alzheimer’s disease; DLB, dementia with Lewy bodies.

### 3.4 SCA FDG-PET as a diagnostic tool

The SCA scores generated from FDG-PET scans in the derivation and independent groups are shown in Figure 3. Within the derivation group, patients with AD had significantly lower SCA scores than those with DLB (F_1,32_ = 73.0, p < 0.001; AD 0.279 ± 0.155, DLB 0.747 ± 0.164). ROC curve analysis showed excellent diagnostic performance in the derivation group (AUC 0.98±0.02, DLB sensitivity 93%, AD specificity 95%). A significant group difference was also found in SCA scores between AD and DLB within the independent group (F_1,31_ = 14.3, p = 0.001; AD 0.418 ± 0.169, DLB 0.649 ± 0.178). Using the optimum decision threshold calculated from the derivation sample, relatively good diagnostic performance was achieved (AUC 0.83±0.07, DLB sensitivity 64%, AD specificity 75%). Using the independent group, diagnostic performance using SCA was compared and showed FDG-PET to be superior to HMPAO-SPECT in distinguishing DLB from AD (Δ_AUC_ 0.24, 95% confidence interval 0.04-0.44, p = 0.020).

**Figure 3.**
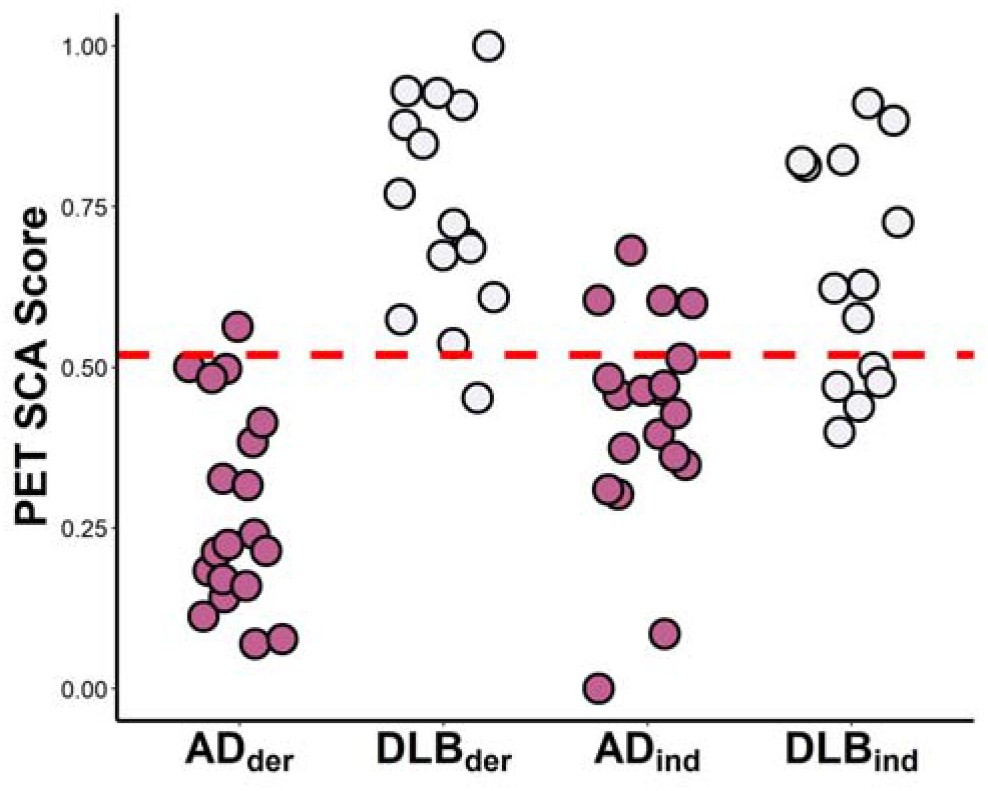
Distribution of SCA scores generated from FDG-PET scans, in the derivation (AD _der_ n=19, DLB_der_ n=15) and independent (AD_ind_ n=19, DLB _ind_ n=14) cohorts. Dotted line indicates separation threshold as calculated from the derivation cohort. Cases above the line were classified as DLB, below as AD. SCA, spatial covariance analysis; AD, Alzheimer’s disease; DLB, dementia with Lewy bodies.

### 3.5 Diagnostic characteristics of SCA, ROI and visual analysis

We then calculated and compared the diagnostic characteristics of visual analysis, ROI analysis and SCA, of FDG-PET and HMPAO-SPECT data, when used to separate DLB from AD in the independent group (Figure 4). We observed good characteristics with ROI analysis of HMPAO-SPECT (AUC 0.82 ± 0.08) and all analyses of FDG-PET (visual AUC 0.82 ± 0.08; ROI AUC 0.84 ± 0.08; SCA AUC 0.83 ± 0.07). Visual interpretation of HMPAO-SPECT was slightly worse (AUC 0.71 ± 0.10), though this was not statistically significant. The lowest performing diagnostic characteristics were seen with SCA of HMPAO-SPECT (AUC 0.59 ± 0.10), though this was only statistically significant when compared to SCA of FDG-PET (reported above). Statistics of all pairwise comparisons are shown in Supplementary tabble 3.

**Figure 4.**
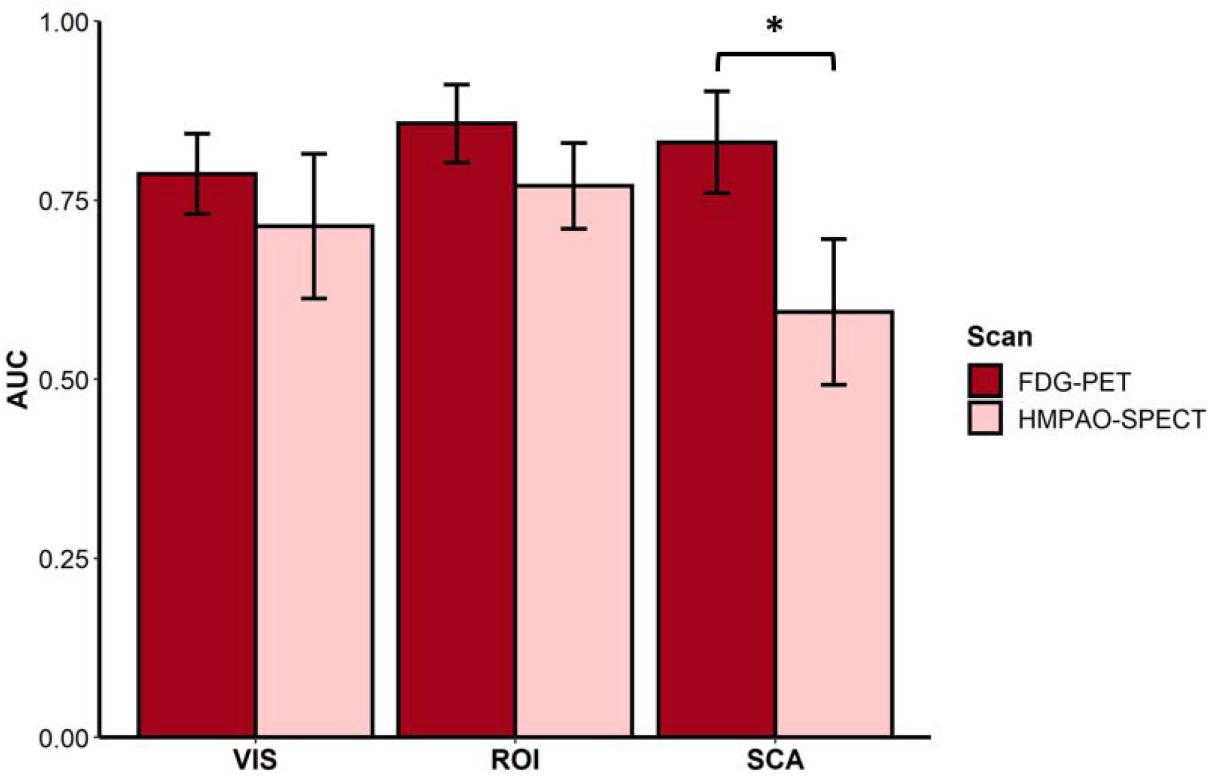
AUCs from ROC analysis of DLB-AD separation with visual analysis (VIS), region of interest (ROI) analysis and spatial covariance analysis (SCA), of FDG-PET and HMPAO-SPECT. ROC analysis performed in the independent group (AD n = 19, DLB n = 14). Error bars denote standard error. AUC, area under ROC curve; ROC, receiver operating characteristics; DLB, dementia with Lewy bodies; AD, Alzheimer’s disease. (p < 0.05, *).

### 3.6 Concordance of quantification methods and visual interpretation in FDG-PET

Finally, as an exploratory analysis we investigated the concordance of visual interpretations of FDG-PET with the ROI method (Figure 5) and SCA (Figure 6). Visually misdiagnosed AD patients (consensus decision of unclear diagnosis or DLB) had higher ROI scores than those who had a correct visual diagnosis (U = 9.000, p = 0.016; 0.415 ± 0.167 [Visually correct mean ± s.d.], 0.619 ± 0.126 [Visually incorrect mean ± s.d.]), and vice versa in the DLB group (U = 1.000, p = 0.003; 0.883 ± 0.079, 0.562 ± 0.212)(Figure 5). Whilst there were broadly similar relationships between visual misdiagnosis and SCA scores (Figure 6), this was not to the same extent as in ROI, and there was no significant difference in group means between the correctly and incorrectly visually diagnosed patients, for either AD or DLB (AD: U = 30.000, p = 0.643; 0.397 ± 0.174, 0.479 ± 0.156. DLB: U = 15.000, p = 0.245; 0.694 ± 0.167, 0.589 ± 0.189). Two (33.3%) of the visually misdiagnosed DLB patients had SCA scores that were strongly indicative of DLB (points circled with dashed red line, Figure 6.A)

**Figure 5.**
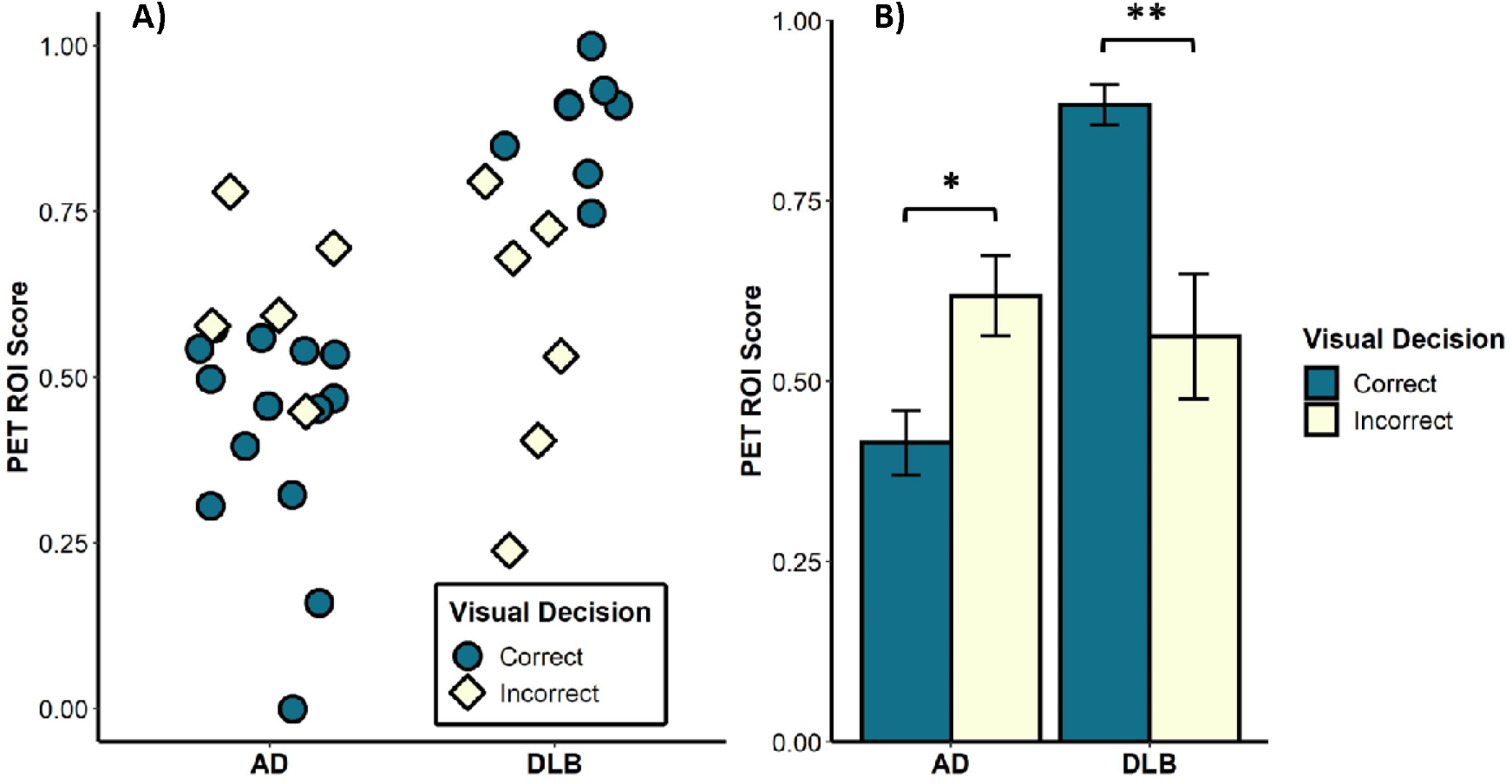
Relationships between ROI quantification and visual interpretation of FDG-PET data. A) FDG-PET ROI scores in AD (n=19) and DLB (n=14) patients, data coloured according to accuracy of visual diagnosis. B) Mean scaled ROI scores in AD and DLB patients, grouped as per accuracy of visual decision. ROI, region of interest; AD, Alzheimer’s disease; DLB, dementia with Lewy bodies. (p < 0.05, * ; p < 0.01, **).

**Figure 6.**
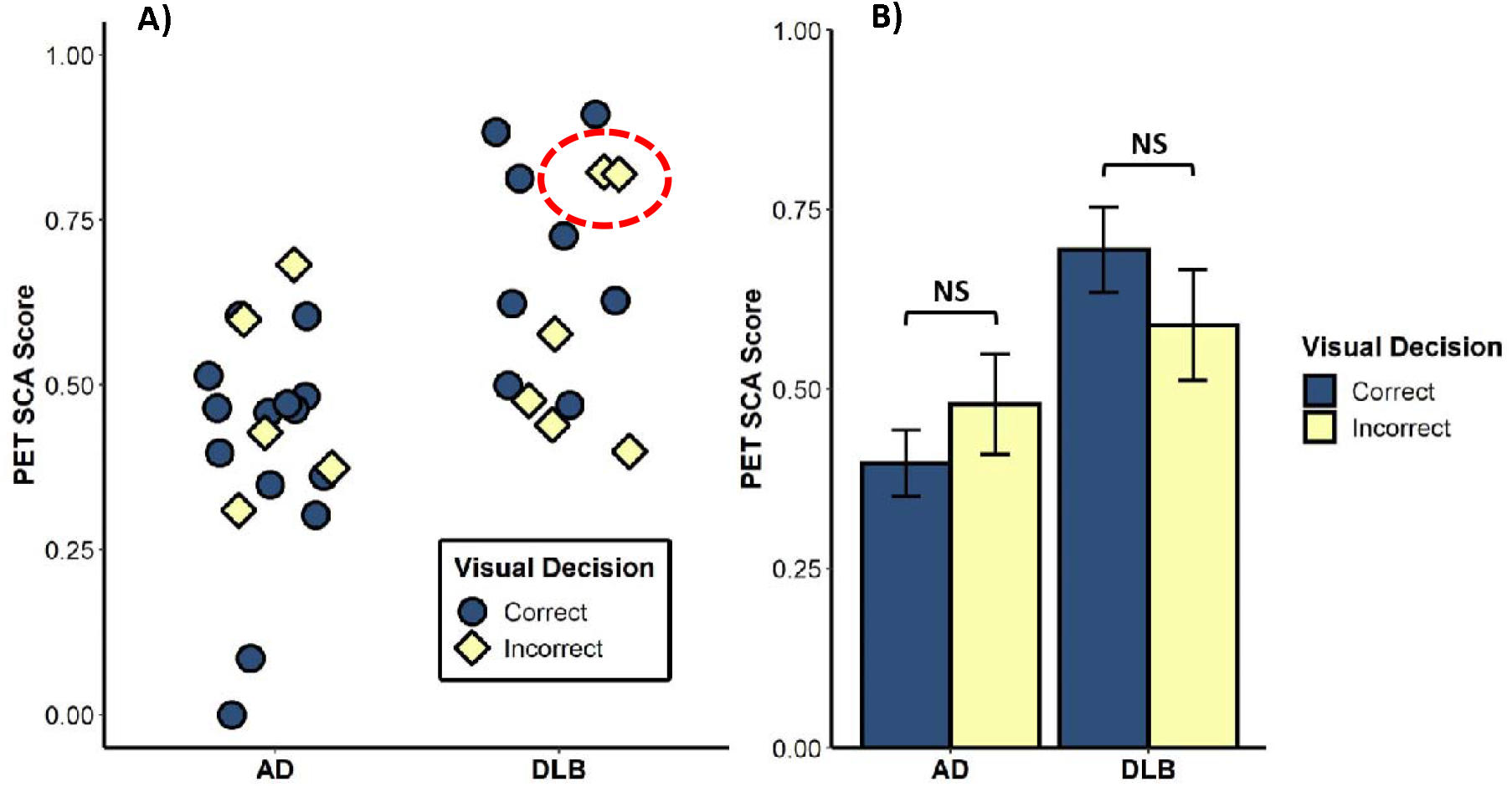
Relationships between SCA quantification and visual interpretation of FDG-PET data. A) FDG-PET SCA scores in AD (n=19) and DLB (n=14) patients, data coloured according to accuracy of visual diagnosis. Dashed red circle indicates the two visually misdiagnosed DLB patients who had SCA’s that were strongly indicative of DLB. B) Mean scaled SSF scores in AD and DLB patients, grouped as per accuracy of visual decision. SCA, spatial covariance analysis; AD, Alzheimer’s disease; DLB, dementia with Lewy bodies. (p > 0.05, not significant (NS)).

Using data from the derivation group, we visually identified the ROI score cut-off (0.671, not shown) that gave optimal DLB-AD separation and forward applied this to ROI scores in the independent group, to give an ROI diagnosis decision for each of those scans (DLB vs. not DLB). When comparing concordance of visual and ROI diagnosis decisions, we found both Cohen’s kappa statistic and percentage agreement to be high (k = 0.669, 95% CI 0.409-0.930: percentage agreement 84.8%). We observed lower Cohen’s kappa (k = 0.405, 95% CI 0.086-0.724) and percentage agreement (72.7%) when comparing SCA and visual diagnosis decisions (DLB vs. not DLB; SCA decision from optimum threshold in section 3.4). Due to the poor diagnostic characteristics of SCA of HMPAO-SPECT, we did not compare measures of concordance between the two quantitative methods and visual analysis with this imaging modality.

## 3. Discussion

We found superior diagnostic characteristics for AD-DLB separation with SCA FDG-PET, compared to SCA HMPAO-SPECT, in both derivation and independent samples. With a derivation group of 19 AD and 15 DLB participants, SCA FDG-PET demonstrated similar diagnostic characteristics to consensus visual interpretation and univariate ROI analysis of FDG-PET scans. We observed weak concordance of SCA FDG-PET with visual interpretations.

Previous studies on visual and univariate quantification analysis of FDG-PET and perfusion SPECT scans have demonstrated in DLB posterior hypoperfusion/hypometabolism with preservation of the posterior cingulate (the cingulate island sign (CIS));^22, 39^ in AD, reductions occur in tracer uptake to temporoparietal regions.^25, 40^ Examination of our FDG-PET SCP reveals a degree of similarity to these visual and univariate signatures, with DLB being distinguished, in part, by decreased concomitant uptake to the occipital lobe and with the CIS clearly visible at the group level. These features are less clear in the HMPAO-SPECT SCP, which may in turn contribute to its poor diagnostic characteristics when applied to the independent group.

We found that in both HMPAO-SPECT and FDG-PET, SCA significantly differentiated the derivation group into AD and DLB. However, when forward applied to the independent group, the diagnostic characteristics for HMPAO-SPECT were poor. SCA of FDG-PET maintained good diagnostic characteristics over the independent group and was significantly better than SCA HMPAO-SPECT. This finding is in accordance with previous work on visual interpretation and univariate analysis, where FDG-PET has been demonstrated as superior to HMPAO-SPECT for the differential diagnosis of AD and DLB.^2, 40^ This is likely the result of well-established factors, including the higher resolution of FDG-PET images compared to SPECT, less noise in the former, and the available PET radioisotopes providing a better surrogate for neuronal activity.^15^ Given the present data, it is likely that FDG-PET would remain the modality of choice over perfusion SPECT for DLB diagnosis, even if multivariate approaches were used clinically.

The poor results shown here for SCA of HMPAO-SPECT (AUC 0.59±0.10) contrast with our previous study, which found very good diagnostic characteristics (AUC 0.86±0.05).^19^ Previously we had used a different dataset (AD n=40, DLB n=26) to derive the HMPAO-SPECT SCP, which was then forward-applied to the entire dataset used in this study (AD n=38, DLB n=29). With the overlap in patients used, the most notable difference is the number of participants. SCA uses the derivation group to generate the optimal separating SCP; theoretically, the larger the derivation group, the smaller the influence of individual variations on the produced SCP, and therefore as derivation group increases in size the SCP better reflects true differences across disease subgroups. Therefore, the smaller numbers in the present study for the HMPAO-SPECT SCP represent a challenge. Another study, using covariance FDG-PET to separate AD patients from healthy controls, found that the derived optimal Z-map varied depending on the composition of derivation groups of 40 subjects,^25^ indicating that larger derivation groups are required to generate the optimal SCP. Given the evidence presented here, that SCA performs significantly better with FDG-PET as opposed to HMPAO-SPECT, and that we can already obtain good AD-DLB diagnostic characteristics with an FDG-PET SCP derived from a modest number of patients (n=29), future studies should explore how far the accuracy of SCA FDG-PET could be further improved with larger derivation groups.

We found that visually analysed FDG-PET, SCA FDG-PET, ROI FDG-PET and ROI HMPAO-SPECT had similar diagnostic characteristics, with visually analysed HMPAO-SPECT being marginally (but not significantly) worse. Given the firm basis for the superiority of FDG-PET,^15, 20^ the relatively strong performances of ROI and visually analysed HMPAO-SPECT, in our comparatively small number of patients, should be taken with some degree of caution. For the FDG-PET data, we found no analytical method to be superior. Despite their inherent advantages of improved standardisation and speed, it is unlikely that quantification methods will become the primary means of analysing neuroimages unless they are shown to consistently outperform visual interpretations. As noted previously, and unlike the ROI method, it is likely that the diagnostic characteristics of SCA FDG-PET for DLB would further improve with larger derivation groups, though it remains unclear as to whether this would be to the point of outperforming visual analysis of perfusion or metabolic scans, or indeed ^123^I-FP-CIT SPECT.

It seems more probable that any clinical application of SCA, or another multivariate method, would be as an adjunct to rather than as a replacement of visual interpretation. As shown in Figure 6.A, comfortable DLB-AD decisions for visual interpreters are not necessarily reflected by correspondingly extreme SCA scores, and vice versa. The potential benefits of this are highlighted in our study, by SCA FDG-PET being able to correctly identify a third of the visually mislabelled DLB patients. Whilst this is a small study, that will need to be replicated, it does demonstrate that SCA of FDG-PET may provide additional diagnostic information compared to visual interpretation alone. SCA could therefore be a financially and computationally inexpensive way of increasing the DLB-sensitivity of FDG-PET. Given that FDG-PET imaging currently finds use in dementia patients with no clinical suspicion of DLB,^26^ this may provide a means of prompting clinical re-evaluation, or consideration of those biomarkers currently considered indicative of DLB.

A notable limitation of this study was our lack of autopsy confirmed cases, and hence our reliance on clinical diagnosis as the gold standard. However, we do note that clinical diagnosis (in research settings) has a high rate of concordance with autopsy.^41^ Another limitation was our small dataset, which may have rendered some analyses underpowered, particularly when we compared SCA diagnostic characteristics with those of visual and ROI analysis. However, this did not affect our principal finding, which was that SCA was superior in FDG-PET over HMPAO-SPECT. Finally, given that the dataset we used only included AD and DLB patients, it would not necessarily be reflective of the clinical population, where healthy patients and other dementia subtypes would also be present. Future studies could investigate SCA of FDG-PET as a means of delineating DLB from this patient group.

In conclusion, SCA FDG-PET significantly outperformed SCA HMPAO-SPECT in the AD-DLB differential diagnosis. Increasing the derivation group size would further improve the diagnostic characteristics of SCA of FDG-PET for DLB diagnosis, but the benefits of this remain unclear and would need further evaluation. Unlike the ROI method, we found that there was only weak concordance between SCA and visual interpretation of FDG-PET data. This provides evidence that additional diagnostic utility can be gained by combining SCA and visual analysis, as supported by our observation that SCA can feasibly identify DLB patients missed by visual interpretation, potentially with no corresponding loss in AD-specificity. Future work should look at the relationship between SCA FDG-PET diagnostic characteristics and derivation group size, with an emphasis on the potential use as an adjunct to visual interpretation; ideally, these studies should assess diagnostic accuracy against pathological findings.

## Data Availability

Data used in the present study are not routinely available.

## Acknowledgements

This paper uses data from independent research funded by the National Institute for Health Research (NIHR) under its Research for Patient Benefit (RfPB) Programme (Grant Reference Number PB-PG-1207-13105). Data collection was supported by the NIHR Newcastle Biomedical Research Centre and Lewybody Dementia Biomedical Research Unit based at Newcastle upon Tyne Hospitals NHS Foundation Trust and Newcastle University, and the NIHR Biomedical Research Centre and Biomedical Research Unit in Dementia based at Cambridge University Hospitals NHS Foundation Trust and the University of Cambridge.

JOB is supported by the NIHR Cambridge Biomedical Centre and the Cambridge Centre for Parkinson’s Plus.

We thank the Dementia and Neurodegenerative Diseases Research Network (DeNDRoN) for support with clinical recruitment.

## References

1. Hogan DB, Fiest KM, Roberts JI, et al. The Prevalence and Incidence of Dementia with Lewy Bodies: a Systematic Review. Can J Neurol Sci 2016; 43 Suppl 1: S83–95. 2016/06/17. DOI: 10.1017/cjn.2016.2.

2. McKeith IG, Boeve BF, Dickson DW, et al. Diagnosis and management of dementia with Lewy bodies: Fourth consensus report of the DLB Consortium. Neurology 2017; 89: 88–100. 2017/06/09. DOI: 10.1212/WNL.0000000000004058.

3. McKeith IG, Dickson DW, Lowe J, et al. Diagnosis and management of dementia with Lewy bodies: third report of the DLB Consortium. Neurology 2005; 65: 1863–1872. 2005/10/21. DOI: 10.1212/01.wnl.0000187889.17253.b1.

4. Rizzo G, Arcuti S, Copetti M, et al. Accuracy of clinical diagnosis of dementia with Lewy bodies: a systematic review and meta-analysis. J Neurol Neurosurg Psychiatry 2018; 89: 358–366. 2017/10/17. DOI: 10.1136/jnnp-2017-316844.

5. Barnes LL, Leurgans S, Aggarwal NT, et al. Mixed pathology is more likely in black than white decedents with Alzheimer dementia. Neurology 2015; 85: 528–534. 2015/07/17. DOI: 10.1212/WNL.0000000000001834.

6. Walker L, McAleese KE, Thomas AJ, et al. Neuropathologically mixed Alzheimer’s and Lewy body disease: burden of pathological protein aggregates differs between clinical phenotypes. Acta Neuropathol 2015; 129: 729–748. 2015/03/12. DOI: 10.1007/s00401-015-1406-3.

7. Taylor JP, McKeith IG, Burn DJ, et al. New evidence on the management of Lewy body dementia. Lancet Neurol 2019 2019/09/15. DOI: 10.1016/S1474-4422(19)30153-X.

8. Galvin JE. Improving the Clinical Detection of Lewy Body Dementia with the Lewy Body Composite Risk Score. Alzheimers Dement (Amst) 2015; 1: 316–324. 2015/09/26. DOI: 10.1016/j.dadm.2015.05.004.

9. Surendranathan A and O’Brien JT. Clinical imaging in dementia with Lewy bodies. Evid Based Ment Health 2018; 21: 61–65. 2018/04/01. DOI: 10.1136/eb-2017-102848.

10. Papathanasiou ND, Boutsiadis A, Dickson J, et al. Diagnostic accuracy of (1)(2)(3)I-FP-CIT (DaTSCAN) in dementia with Lewy bodies: a meta-analysis of published studies. Parkinsonism Relat Disord 2012; 18: 225–229. 2011/10/07. DOI: 10.1016/j.parkreldis.2011.09.015.

11. NICE. Dementia: assessment, management and support for people living with dementia and their carers, https://www.nice.org.uk/guidance/ng97/chapter/Recommendations#diagnosis (2018, accessed 23/06/2021).

12. McKeith I, O’Brien J, Walker Z, et al. Sensitivity and specificity of dopamine transporter imaging with 123I-FP-CIT SPECT in dementia with Lewy bodies: a phase III, multicentre study. Lancet Neurol 2007; 6: 305–313. 2007/03/17. DOI: 10.1016/S1474-4422(07)70057-1.

13. Yoshita M, Arai H, Arai H, et al. Diagnostic accuracy of 123I-meta-iodobenzylguanidine myocardial scintigraphy in dementia with Lewy bodies: a multicenter study. PLoS One 2015; 10: e0120540. 2015/03/21. DOI: 10.1371/journal.pone.0120540.

14. Yousaf T, Dervenoulas G, Valkimadi PE, et al. Neuroimaging in Lewy body dementia. J Neurol 2019; 266: 1–26. 2018/05/16. DOI: 10.1007/s00415-018-8892-x.

15. Laforce R, Jr., Soucy JP, Sellami L, et al. Molecular imaging in dementia: Past, present, and future. Alzheimers Dement 2018; 14: 1522–1552. 2018/07/22. DOI: 10.1016/j.jalz.2018.06.2855.

16. Ferrando R and Damian A. Brain SPECT as a Biomarker of Neurodegeneration in Dementia in the Era of Molecular Imaging: Still a Valid Option? Front Neurol 2021; 12: 629442. 2021/05/28. DOI: 10.3389/fneur.2021.629442.

17. Kemp PM, Hoffmann SA, Tossici-Bolt L, et al. Limitations of the HMPAO SPECT appearances of occipital lobe perfusion in the differential diagnosis of dementia with Lewy bodies. Nucl Med Commun 2007; 28: 451–456. 2007/04/27. DOI: 10.1097/MNM.0b013e328155d143.

18. Caminiti SP, Sala A, Iaccarino L, et al. Brain glucose metabolism in Lewy body dementia: implications for diagnostic criteria. Alzheimers Res Ther 2019; 11: 20. 2019/02/25. DOI: 10.1186/s13195-019-0473-4.

19. Colloby SJ, Taylor JP, Davison CM, et al. Multivariate spatial covariance analysis of 99mTc-exametazime SPECT images in dementia with Lewy bodies and Alzheimer’s disease: utility in differential diagnosis. J Cereb Blood Flow Metab 2013; 33: 612–618. 2013/01/31. DOI: 10.1038/jcbfm.2013.2.

20. O’Brien JT, Firbank MJ, Davison C, et al. 18F-FDG PET and perfusion SPECT in the diagnosis of Alzheimer and Lewy body dementias. J Nucl Med 2014; 55: 1959–1965. 2014/12/03. DOI: 10.2967/jnumed.114.143347.

21. Kantarci K, Lowe VJ, Boeve BF, et al. Multimodality imaging characteristics of dementia with Lewy bodies. Neurobiol Aging 2012; 33: 2091–2105. 2011/10/25. DOI: 10.1016/j.neurobiolaging.2011.09.024.

22. Lim SM, Katsifis A, Villemagne VL, et al. The 18F-FDG PET cingulate island sign and comparison to 123I-beta-CIT SPECT for diagnosis of dementia with Lewy bodies. J Nucl Med 2009; 50: 1638–1645. 2009/09/18. DOI: 10.2967/jnumed.109.065870.

23. Scarmeas N, Habeck CG, Zarahn E, et al. Covariance PET patterns in early Alzheimer’s disease and subjects with cognitive impairment but no dementia: utility in group discrimination and correlations with functional performance. Neuroimage 2004; 23: 35–45. 2004/08/25. DOI: 10.1016/j.neuroimage.2004.04.032.

24. Colloby SJ, Taylor JP, Firbank MJ, et al. Covariance 99mTc-exametazime SPECT patterns in Alzheimer’s disease and dementia with Lewy bodies: utility in differential diagnosis. J Geriatr Psychiatry Neurol 2010; 23: 54–62. 2009/12/24. DOI: 10.1177/0891988709355272.

25. Habeck C, Foster NL, Perneczky R, et al. Multivariate and univariate neuroimaging biomarkers of Alzheimer’s disease. Neuroimage 2008; 40: 1503–1515. 2008/03/18. DOI: 10.1016/j.neuroimage.2008.01.056.

26. Dementia: assessment, management and support for people living with dementia and their carers, https://www.nice.org.uk/guidance/ng97/chapter/Recommendations#diagnosis (2018, accessed 1st November 2019).

27. McKhann G, Drachman D, Folstein M, et al. Clinical diagnosis of Alzheimer’s disease: report of the NINCDS-ADRDA Work Group under the auspices of Department of Health and Human Services Task Force on Alzheimer’s Disease. Neurology 1984; 34: 939–944. 1984/07/01. DOI: 10.1212/wnl.34.7.939.

28. Roth M HF, Mountjoy CQ, Tym E. CAMDEX-R: Revised Cambridge Examination for Mental Disorders of the Elderly. Cambridge, UK: Cambridge University Press, 1999.

29. Arevalo-Rodriguez I, Smailagic N, Roque IFM, et al. Mini-Mental State Examination (MMSE) for the detection of Alzheimer’s disease and other dementias in people with mild cognitive impairment (MCI). Cochrane Database Syst Rev 2015: CD010783. 2015/03/06. DOI: 10.1002/14651858.CD010783.pub2.

30. Post B, Merkus MP, de Bie RM, et al. Unified Parkinson’s disease rating scale motor examination: are ratings of nurses, residents in neurology, and movement disorders specialists interchangeable? Mov Disord 2005; 20: 1577–1584. 2005/08/24. DOI: 10.1002/mds.20640.

31. Habeck C, Krakauer JW, Ghez C, et al. A new approach to spatial covariance modeling of functional brain imaging data: ordinal trend analysis. Neural Comput 2005; 17: 1602–1645. 2005/05/20. DOI: 10.1162/0899766053723023.

32. Habeck C and Stern Y. Multivariate data analysis for neuroimaging data: overview and application to Alzheimer’s disease. Cell Biochem Biophys 2010; 58: 53–67. 2010/07/27. DOI: 10.1007/s12013-010-9093-0.

33. Tzourio-Mazoyer N, Landeau B, Papathanassiou D, et al. Automated anatomical labeling of activations in SPM using a macroscopic anatomical parcellation of the MNI MRI single-subject brain. Neuroimage 2002; 15: 273–289. 2002/01/05. DOI: 10.1006/nimg.2001.0978.

34. Colloby SJ, Fenwick JD, Williams ED, et al. A comparison of (99m)Tc-HMPAO SPET changes in dementia with Lewy bodies and Alzheimer’s disease using statistical parametric mapping. Eur J Nucl Med Mol Imaging 2002; 29: 615–622. 2002/04/27. DOI: 10.1007/s00259-002-0778-5.

35. Zweig MH and Campbell G. Receiver-operating characteristic (ROC) plots: a fundamental evaluation tool in clinical medicine. Clin Chem 1993; 39: 561–577. 1993/04/01.

36. DeLong ER, DeLong DM and Clarke-Pearson DL. Comparing the areas under two or more correlated receiver operating characteristic curves: a nonparametric approach. Biometrics 1988; 44: 837–845. 1988/09/01.

37. McHugh ML. Interrater reliability: the kappa statistic. Biochem Med (Zagreb) 2012; 22: 276–282. 2012/10/25.

38. Ranganathan P, Pramesh CS and Aggarwal R. Common pitfalls in statistical analysis: Measures of agreement. Perspect Clin Res 2017; 8: 187–191. 2017/11/08. DOI: 10.4103/picr.PICR_123_17.

39. Graff-Radford J, Murray ME, Lowe VJ, et al. Dementia with Lewy bodies: basis of cingulate island sign. Neurology 2014; 83: 801–809. 2014/07/25. DOI: 10.1212/WNL.0000000000000734.

40. McKhann GM, Knopman DS, Chertkow H, et al. The diagnosis of dementia due to Alzheimer’s disease: recommendations from the National Institute on Aging-Alzheimer’s Association workgroups on diagnostic guidelines for Alzheimer’s disease. Alzheimers Dement 2011; 7: 263–269. 2011/04/26. DOI: 10.1016/j.jalz.2011.03.005.

41. Snowden JS, Thompson JC, Stopford CL, et al. The clinical diagnosis of early-onset dementias: diagnostic accuracy and clinicopathological relationships. Brain 2011; 134: 2478–2492. 2011/08/16. DOI: 10.1093/brain/awr189.

