## Supplemental Materials for "Spatial covariance analysis of FDG-PET and HMPAO-SPECT for the differential diagnosis of dementia with Lewy bodies and Alzheimer’s disease"

**Supplementary Materials**

| Hemisphere | MNI coordinates | Region | Z score |
| --- | --- | --- | --- |
| R | 34, -76, -44 | Posterior cerebellum | 1.7 |
| L | -28, -74, -44 | Posterior cerebellum | 1.7 |
| R | 12, -22, 58 | Medial frontal gyrus | 2.2 |
| L | 0, -62, -38 | Cerebellar tonsil | 1.8 |
| R | 50, -16, -32 | Inferior temporal gyrus | 1.7 |
| L | -30, 16, -40 | Superior temporal gyrus | 1.7 |
| L | -34, -8, -32 | Uncus | 2.1 |
| L | 0, -58, -32 | Anterior cerebellum | 1.8 |
| L | -36, -6, -26 | Amygdala | 2.2 |
| L | -38, -10, -20 | Hippocampus | 2.0 |
| L | -8, -52, 54 | Precuneus | 2.3 |
| R | 36, 58, -10 | Superior frontal gyrus | -1.7 |
| L | -8, 42, 52 | Superior frontal gyrus | -1.7 |
| R | 44, -84, 4 | Inferior occipital gyrus | -1.8 |
| L | -42, -88, 4 | Inferior occipital gyrus | -1.9 |
| R | 54, -2, 52 | Precentral gyrus | -1.7 |
| L | -40, -12, 32 | Precentral gyrus | -1.9 |
| L | -14, -94, -14 | Lingual gyrus | -1.9 |
| L | -44, -84, 14 | Middle occipital gyrus | -1.8 |

**Supplementary Table 1**. Location of regions contributing to the derivation HMPAO-SPECT pattern distinguishing DLB from AD. MNI, Montreal Neurological Institute.

| Hemisphere | MNI coordinates | Region | Z score |
| --- | --- | --- | --- |
| R | 40, -22, -18 | Hippocampus | 2.1 |
| L | -30, -20, -18 | Hippocampus | 2.0 |
| R | 26, -6, -30 | Amygdala | 1.8 |
| L | -30, -2, -30 | Amygdala | 1.9 |
| L | -34, 18, -46 | Superior temporal gyrus | 2.2 |
| L | -64, -20, -22 | Inferior temporal gyrus | 1.8 |
| L | -46, 8, -40 | Middle temporal gyrus | 1.8 |
| L | -34, 34, -18 | Inferior frontal gyrus | 1.7 |
| L | 0, -30, 38 | Posterior cingulate | 2.4 |
| L | 0 -52, -30 | Anterior cerebellum | 2.0 |
| R | 20, -44, 38 | Precuneus | -2.0 |
| L | -18, -46, 38 | Precuneus | -2.1 |
| R | 68, -20, 24 | Inferior parietal lobe | -1.9 |
| L | -46, -42, 58 | Inferior parietal lobe | -1.7 |
| R | 52, 28, 24 | Middle frontal gyrus | -1.8 |
| L | -34, 18, 58 | Middle frontal gyrus | -1.8 |
| R | 14, 34, 50 | Superior frontal gyrus | -1.8 |
| R | 46, 38, 6 | Inferior frontal gyrus | -1.8 |
| L | -14, -68, 0 | Lingual gyrus | -2.0 |

**Supplementary Table 2**. Location of regions contributing to the derivation FDG-PET pattern distinguishing DLB from AD. MNI, Montreal Neurological Institute.

| Comparison | AUC difference | 95% CI | P |
| --- | --- | --- | --- |
| FDG-PET visual -  HMPAO-SPECT visual | .102 | -0.32-0.235 | 0.136 |
| FDG-PET visual -  FDG-PET SCA | -.015 | -0.200-0.170 | 0.874 |
| FDG-PET visual -  FDG-PET ROI | -.019 | -0.108-0.070 | 0.679 |
| FDG-PET visual -  HMPAO-SPECT SCA | .222 | -0.38-0.481 | 0.094 |
| FDG-PET visual -  HMPAO-SPECT ROI | -.004 | -0.069-0.062 | 0.910 |
| HMPAO-SPECT visual - FDG-PET SCA | -.117 | -0.332-0.098 | 0.288 |
| HMPAO-SPECT visual - FDG-PET ROI | -.120 | -0.281-0.040 | 0.142 |
| HMPAO-SPECT visual - HMPAO-SPECT SCA | .120 | -0.176-0.417 | 0.426 |
| HMPAO-SPECT visual - HMPAO-SPECT ROI | -.105 | -0.236-0.026 | 0.115 |
| FDG-PET SCA -  FDG-PET ROI | -.004 | -0.177-0.170 | 0.996 |
| FDG-PET SCA -  HMPAO-SPECT SCA | .237 | 0.037-0.436 | 0.020 * |
| FDG-PET SCA -  HMPAO-SPECT ROI | .011 | -0.190-0.213 | 0.913 |
| FDG-PET ROI -  HMPAO-SPECT SCA | .241 | -0.21-0.502 | 0.072 |
| FDG-PET ROI -  HMPAO-SPECT ROI | .015 | -0.074-0.104 | 0.742 |
| HMPAO-SPECT SCA - HMPAO-SPECT ROI | -.226 | -0.491-0.040 | 0.095 |

**Supplementary Table 3**. Pairwise comparisons in the DLB-AD diagnostic characteristics of spatial covariance analysis (SCA), region of interest (ROI) analysis and visual ratings of FDG-PET and HMPAO-SPECT scans, in the independent group (n=34). AUC, area under receiver operating characteristics curve; CI, confidence interval; DLB, dementia with Lewy bodies; AD, Alzheimer’s disease. (p < 0.05, *).
